# Effectiveness of Isolation Policies in Schools: Evidence from a Mathematical Model of Influenza and COVID-19

**DOI:** 10.1101/2020.03.26.20044750

**Authors:** Adam A. C. Burns, Alexander Gutfraind

**Affiliations:** Department of Medicine, Loyola University Medical Center, Maywood, IL, USA; School of Public Health, University of Illinois at Chicago, Chicago, IL, USA

## Abstract

**Background:** Non-pharmaceutical interventions such as social distancing, school closures and travel restrictions are often implemented to control outbreaks of infectious diseases. For influenza in schools, the Center of Disease Control (CDC) recommends that febrile students remain isolated at home until they have been fever-free for at least one day and a related policy is recommended for SARS-CoV2 (COVID-19). Other authors proposed using a school week of four or fewer days of in-person instruction for all students to reduce transmission. However, there is limited evidence supporting the effectiveness of these interventions.

**Methods:** We introduced a mathematical model of school outbreaks that considers both intervention methods. Our model accounts for the school structure and schedule, as well as the time-progression of fever symptoms and viral shedding. The model was validated on outbreaks of seasonal and pandemic influenza and COVID-19 in schools. It was then used to estimate the outbreak curves and the proportion of the population infected (attack rate) under the proposed interventions.

**Results:** For influenza, the CDC-recommended one day of post-fever isolation can reduce the attack rate by a median (interquartile range) of 29 (13 - 59)%. With two days of post-fever isolation the attack rate could be reduced by 70 (55 - 85)%. Alternatively, shortening the school week to four and three days reduces the attack rate by 73 (64 - 88)% and 93 (91 - 97)%, respectively. For COVID-19, application of post-fever isolation policy was found to be less effective and reduced the attack rate by 10 (5 - 17)% for a two-day isolation policy and by 14 (5 - 26)% for 14 days. A four-day school week would reduce the median attack rate in a COVID-19 outbreak by 57 (52 - 64)%, while a three-day school week would reduce it by 81 (79 - 83)%. In both infections, shortening the school week significantly reduced the duration of outbreaks.

**Conclusions:** Shortening the school week could be an important tool for controlling influenza and COVID-19 in schools and similar settings. Additionally, the CDC-recommended post-fever isolation policy for influenza could be enhanced by requiring two days of isolation instead of one.

## Introduction

Respiratory infections are the leading cause of death in low- or middle-income countries and account for an estimated four million deaths annually^1^. For rapidly emerging outbreaks such as novel strains of influenza or SARS-CoV-2, pharmaceutical measures may be unavailable or ineffective, in which case non-pharmaceutical intervention measures (NPIs) may be the first line of response against infection^2,3^. However, when the World Health Organization (WHO) systematically reviewed all studies supporting NPIs for controlling pandemic influenza, many of the available NPIs lacked sufficient evidence of effectiveness^4^ and it was unclear whether these measures could be effective at controlling COVID-19 (the disease caused by the SARS-CoV-2 virus).

Our study directly addresses this gap by using a computational model to examine the impact of several NPIs, focusing on a shortened school week and symptom-based isolation policies. The shortened school week policy involves closure of the school for additional days to extend the weekend (e.g. closure every Thursday and Friday)^5,6^, thus creating a period of additional physical isolation between the students, and possibly conducting at-home learning on those days. The symptom-based isolation policy involves isolating individuals at the onset and for the duration of fever symptoms, normally followed by additional days of isolation.

For controlling pandemic and seasonal influenza outbreaks, the symptom-based NPI with one day of post-fever isolation is currently recommended by the US Centers of Disease Control and Prevention (CDC)^7,8^, and is referred to as fever absenteeism or a return-to-school policy^9^. The buffer period of one day reduces transmission from infectious students in situations where their fever symptoms temporarily subside or when they are shedding virus at the end of the course of illness^10^.

For COVID-19 outbreaks, there are currently divergent recommendations from public health authorities, and there have been several changes in guidelines. For symptomatic patients, the WHO currently recommends 10 days of isolation after symptom onset, plus at least 3 additional days without symptoms, while for asymptomatic patients it recommends 10 days after positive test for SARS-CoV-2^11^. The CDC recommends that patients with COVID-19 symptoms isolate until they have met all three conditions: (1) they have been fever-free for 24 hours without the use of fever-reducing medications, (2) it has been 10 days since the onset of their symptoms and (3) their COVID-19 symptoms have been improving^12^. Finally, the Swedish Public Health Agency (SPHA), which previously required two days free of symptoms, currently gives the schools control in establishing their own isolation policies^13^.

To evaluate the various NPIs, our model computationally simulates outbreaks of influenza and COVID-19 in school settings, and then looks at the effect of isolation policies on the attack rate (i.e. the proportion of the student population infected over the duration of the outbreak) and the outbreak curve (i.e. prevalence of infected students at each day of the outbreak). Our work builds upon previous studies that have applied mathematical models to school-based influenza transmission^14–17^. The policy of symptom monitoring, which isolates contacts after onset of symptoms, has been examined computationally and shown to be sufficient for controlling certain outbreaks^18^. Several studies also modeled non-pharmaceutical interventions such as closures but not absenteeism policies^17,19–30^. We retrieved all studies that evaluated isolation policies by using a broad search on PubMed and found that many modeling studies assumed isolation for a fixed interval following diagnosis but not in a symptom-dependent way^16^. We also found several comprehensive computational studies of school closures and isolation of infected students^16,31^, but did not find evaluation of symptom-based isolation policies. The policy of shortening school weeks was recently examined in a modeling study^6^ that found that closures for ten days followed by four days of schooling could be effective in controlling COVID-19, but our model is the first to examine the modification of a standard five-day school schedule.

Despite the promise of post-fever isolation for both COVID-19 and influenza, we hypothesized at the outset of this project that the policy would merely have a small effect in controlling outbreaks. We speculated that the CDC-recommended single day of post-fever isolation might not be enough to achieve a meaningful reduction in influenza transmission. Furthermore, parental non-compliance may make the policy ineffective. As compared to influenza, the policy would be less effective in COVID-19 outbreaks since children have a higher rate of asymptomatic infections^32^. On the other hand, if the policy were proven effective for either outbreak, it is not known whether one day of post-fever isolation is optimal, and the model could help determine if additional days of isolation would be beneficial. Additionally, we hypothesized that the short school-week policy, although disruptive, may be more effective for both infections since it could be easier to enforce than symptom-based isolation. In the case of COVID-19, the policy has the advantage of not being affected by the low rate of symptomatic infections among children^32^.

## Materials and Methods

We use a deterministic compartmental dynamical model known as the Susceptible, Exposed, Infectious, Recovered (SEIR) model that tracks the number of individuals of various cohorts and immunological states for each day during an outbreak (Figure 1). This class of SEIR models has been used extensively to model influenza^14,20,33,34^ and COVID-19^35,36^, and we extended the SEIR framework in order to account for isolation policies.

**Figure 1:**
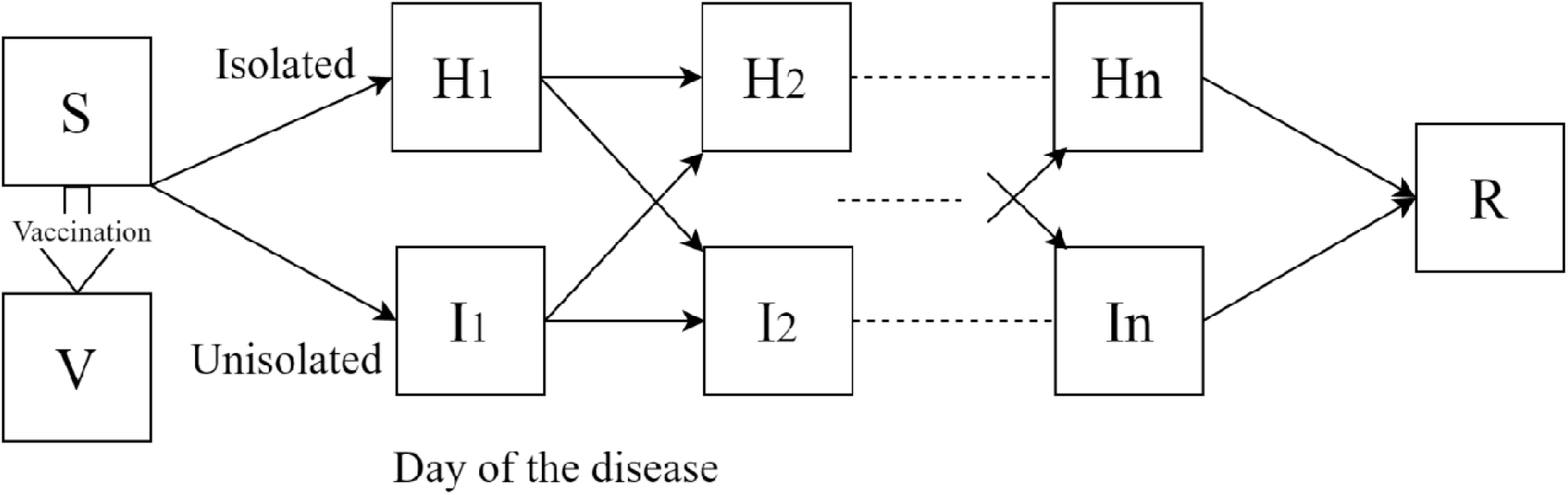
Dynamics of the outbreak model and its major variables. S, susceptible; H, infected isolated; I, infected unisolated; R, recovered; and V, vaccinated or immune (if a vaccine is available). The infected classes (H and I) are stratified to cohorts (shown here is the case of a single cohort), and *n* daily disease stages distinguished by severity of symptoms and viral shedding (9 daily stages of viral shedding for influenza and 32 stages for COVID-19, see Supplementary Article S1). The cohort structure and other parameters are adjustable to model outbreaks in different settings and by different pathogens. Vaccination is available only for some outbreaks and generally has incomplete coverage and efficacy.

Our model is particularly novel as it further stratifies the population by both the day of their infection, their location (isolated at home vs. not isolated in school) and their grade, with students in the same grade generally having closer contacts to peers in the same grade^37^ (see Supplemental Article S1 - Expanded Methods for details). In the model, the day of infection determines the rate of virus shedding and the probability of symptoms, which then influences the likelihood of isolating at home or returning to school. The probability of isolating was also based on the stage of the illness, as well as the isolation policy. The model allowed a vaccine to be received by some students, if the vaccine is available ahead of the outbreak, attaining partial protection against the infection. The model also considered any pre-existing immunity, the rate of symptomatic infections, and school holidays. Policy analysis used the setting of a typical school (6 grades with 70 students each)^38^, but also considered alternative scenarios with larger schools (140 students per grade)

The model was validated on outbreaks of influenza and COVID-19 in schools and shown to match the peak and duration of the outbreak curves, and the overall attack rates of the student population. The validation data was from two outbreaks of pandemic influenza^39,40^, one outbreak of seasonal influenza^41^, and one outbreak of COVID-19^42^. Model parameter ranges were derived from published sources and by calibration to data using a stochastic optimization algorithm. To ensure that the results were robust to uncertainty in parameter values, we then simulated the epidemic 500 times per scenario to account for possible difference between schools and seasons, with normally distributed values for parameters such as the start day in the year, contact rate between cohorts and others, and reported the median and the interquartile ranges. All modeling and statistical analysis used the RStudio Integrated Development for R. RStudio, Inc., Boston, MA. Further details on the model, including the equations and the parameter values, are provided in Supplementary Article S1.

Using the model we considered the effect of two key control policies, fever-based isolation and a shortened school week. For fever-based isolation we evaluated the effect of stricter compliance, which could be attained by remote monitoring, help in maintaining home isolation or penalties for non-compliance. We also considered the effect of increasing the monitoring of symptoms, which could be attained through training of the parents and distribution of free thermometers. We also considered supplemental policies such as subdividing students into small cohorts and enforcing strict quarantines on weekends.

## Results

Our analysis considered the effect of symptom-based isolation and alternative school schedule policies on influenza and SARS-CoV-2 infections. The symptom-based policy is defined as a student remaining in isolation a number of days following a fever whereas the school schedule policy is defined as the number of in-person instruction days students attend weekly. The effect of these policies is summarized in Table 1 and described in detail below.

**Table 1.** Relative effectiveness (%) of isolation policies in schools on the attack rate and outbreak duration compared to the baseline outbreaks without interventions. Median effect (interquartile range). For the policy and baseline scenarios, the effective attack rate and outbreak duration were calculated from the median of a set of 500 simulations for each policy option.

| **Outbreak** | **Policy option** | **Attack rate**<br>**(% decrease)** | **Outbreak duration**<br>**(% decrease)** |
| --- | --- | --- | --- |
| ***Flu*** | One day isolation<br>(CDC guideline) | 29 (13 - 59)% | 1 (-2 – 16)% |
|  | Two day post-fever<br>isolation | 70 (55 - 85)% | 18 (6 – 66)% |
|  | Four day school<br>week | 73 (64 - 88)% | 20 (11 – 55)% |
|  | Three day school<br>week | 93 (91 - 97)% | 99 (82 – 100)% |
| ***COVID-19*** | One day isolation | 7 (5 - 14)% | 1 (1 – 4)% |
|  | Two day post-fever<br>isolation | 10 (5 - 17)% | 1 (1 – 4)% |
|  | 14 day post-fever<br>isolation | 14 (5 - 26)% | 4 (3 – 7)% |
|  | Four day school<br>week | 57 (52 - 64)% | 22 (12 – 26)% |
|  | Three day school<br>week | 81 (79 - 83)% | 46 (33 – 52)% |

### Influenza

We calibrated our model’s parameters to outbreaks of pandemic and seasonal influenza and then calculated each isolation policy 500 times. The different runs varied parameters that normally vary from year to year (e.g. outbreak start day and compliance with policy). In the baseline scenario of pandemic influenza and no isolation policy, our model has a median attack rate of 24.5 (Interquartile Range, IQR: 16.6 - 28.1)%. Implementing a one- and two-day isolation policy, our model predicts a decrease in the attack rate to 17.2 (9.9 - 21.4)% and 7.4 (3.7 - 11.1)%, respectively (Figure 2).

**Figure 2.**
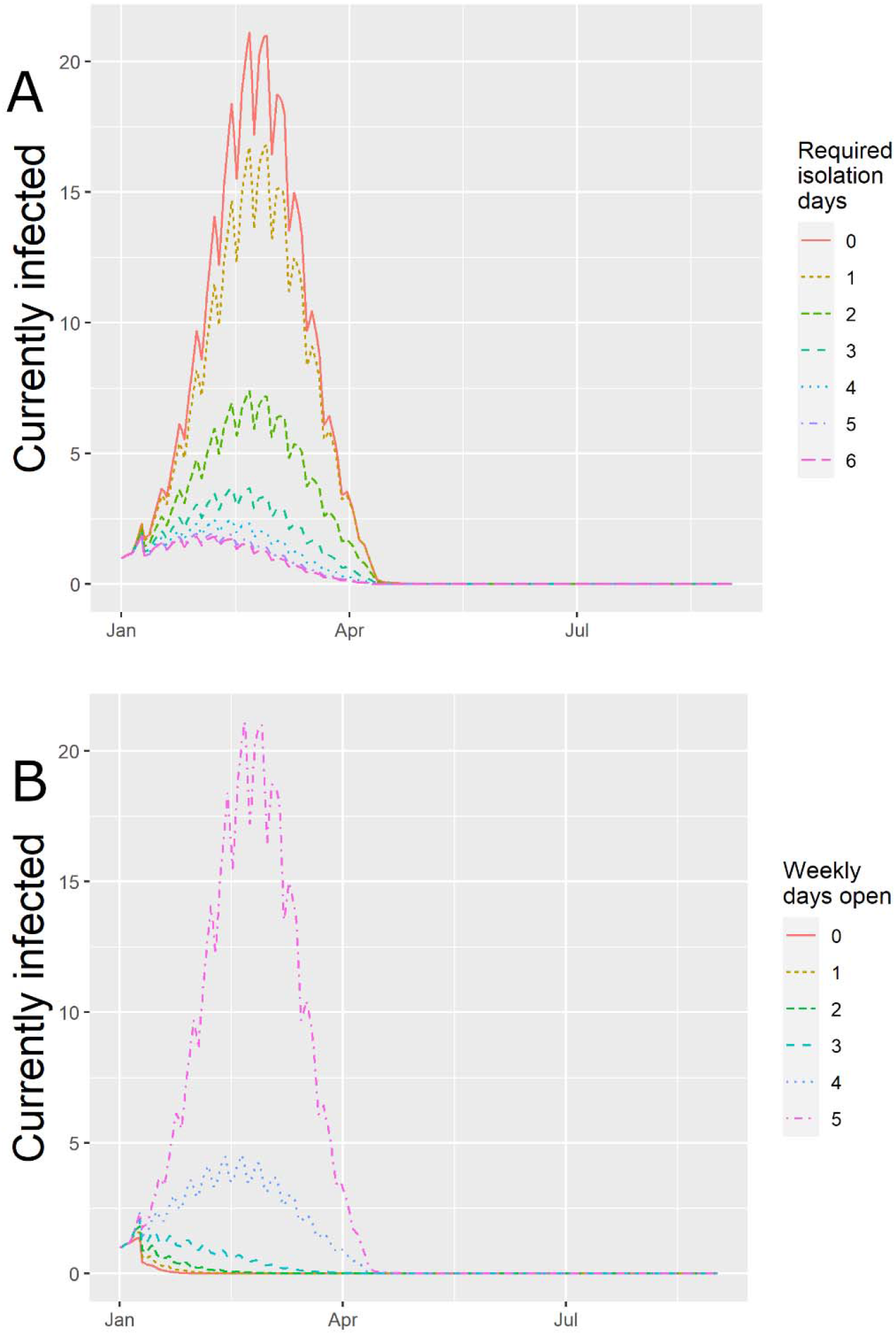
The effect of requiring isolation after the last fever event in a median US school experiencing an outbreak of influenza (A) fever isolation and (B) shortened in-person school week. Vertical axis indicates daily prevalence and ripples are due to weekends and closures. Summer holiday starts June 17 and reduces transmission. Increasing the required days of isolation or shortening the in-person school week reduces the peak infected and the number concurrently infected. Only shortening the in-person school week reduces the duration of the outbreak.

Furthermore, the model predicts that a two-day policy reduces the peak prevalence (i.e. simultaneously infected students) from 20 (13 - 25) to 5 (2 - 8) and the outbreak duration from 82 (78 - 84) to 67 (28 - 77) days. Our model suggests that the two-day policy is still effective even when the student population has pre-existing immunity or has been vaccinated (mean vaccination rate of 80% and efficacy of 50%), reducing the attack rate to 13 (4.2 - 23.8)%.

A policy of a four-day school week gives a 73% reduction in the attack rate from the baseline, to 6.8 (3.3 - 8.8)%, and a three-day school week gives a 93% reduction to 1.8 (0.9 - 2.3) (Figure 2B). The two-day of post-fever isolation and three-day week policies could be combined additively with two days of post-fever isolation to give attack rates of 2.1 (1.0-3.3)% and 0.9 (0.5 - 1.2)%, respectively.

### COVID-19 (SARS-CoV-2)

In our COVID-19 outbreak scenarios, we calibrated the model to a baseline attack rate of 10.0 (8.7 - 11.3)% in 500 simulations. As expected based on the lower rate of symptomatic infections in COVID-19 as compared to influenza, post-fever isolation policy is less effective for COVID-19: 1, 2, and 14 days of isolation yielded attacks rates of 9.4 (8.3 - 10.6)%, 9.2 (8.0 - 10.6)%, and 8.5 (7.4 - 9.7)%, respectively (Figure 3, A). Additionally, our model had a baseline outbreak duration of 138 (135 - 140) days and yielded outbreak durations of 137 (133 - 139), 136 (132 - 139), and 132 (128 - 134) days for 1, 2, and 14 days of post-fever isolation. Thus when compared to the influenza outbreaks, this type of policy was less effective at attack rate or outbreak duration inhibition.

**Figure 3.**
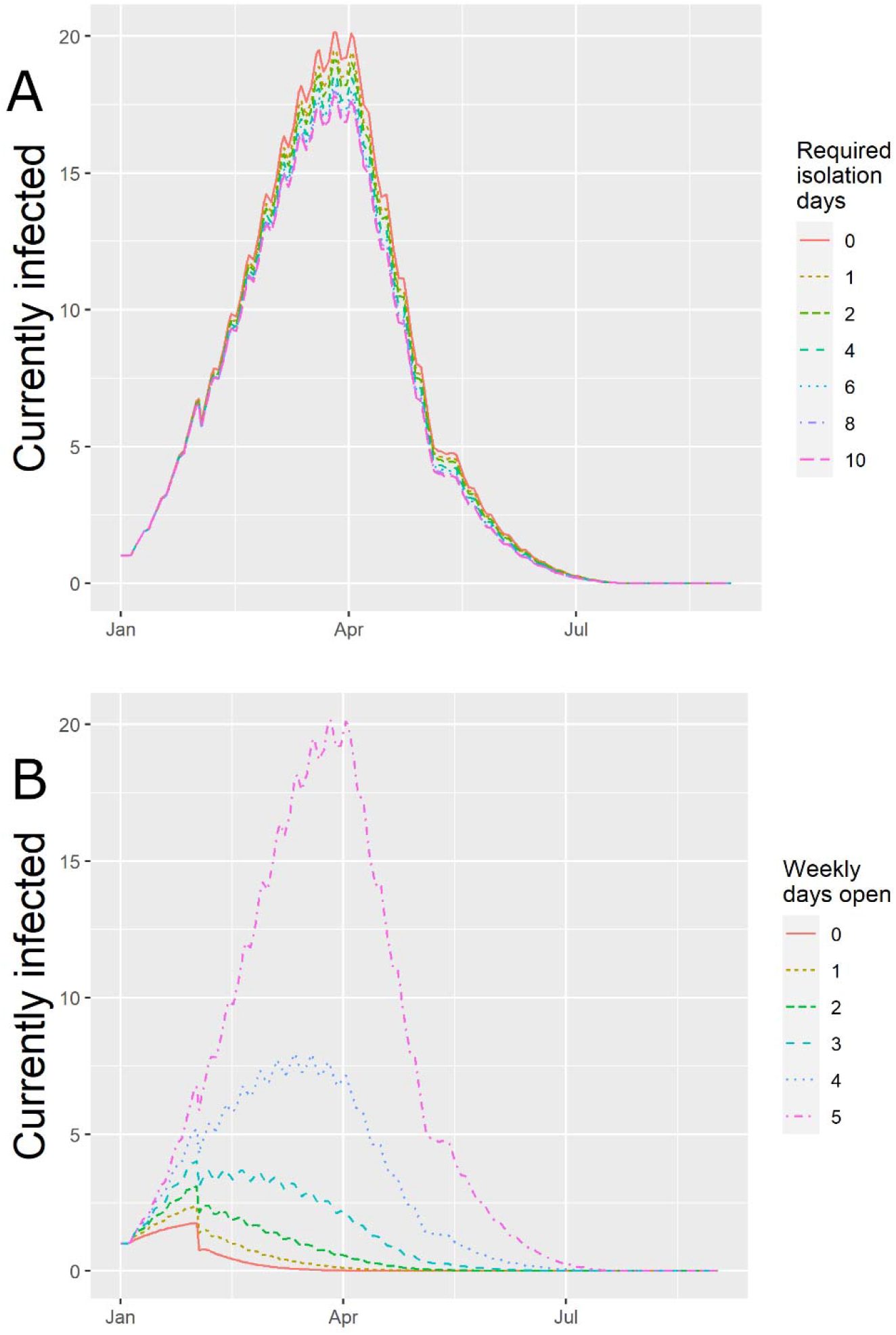
The effect of (A) post-fever isolation and (B) in-person school-week reduction policies on a median US school experiencing an outbreak of COVID-19. Vertical axis indicates daily prevalence as in Fig 2. Increasing the number of post-fever isolation days has little effect on the outbreak. Reducing the number of school days that students physically go to school each week reduces the peak number of infected, the number concurrently infected, and the duration of the outbreak.

Evaluating the policy of shortening the school week, our model found that using four and three days of in-person schooling yields attack rates of 4.4 (3.7 - 4.9)% and 2.0 (1.7 - 2.2)%, respectively (Figure 3, B). When the student population is presumed to be immune^43^ or vaccinated at a rate of 80% and with an efficacy of 70% (Standard Deviation, SD: 20%), the model predicts that the policy is still effective: for the 4- and 3-day school week, the model predicts a median attack rate of 5.1 (1.8-15.2)% and 2.7 (1-6.6) %, respectively.

For both influenza and COVID-19, we examined additional policy options to complement the two main policies of post-fever isolation and shortening the school week. In the case of symptom-based isolation for influenza, measures that increase compliance and symptom monitoring were found to be effective: a 25% increase in attention and compliance reduces the attack rate to 3.4 (1.5 - 5.2)% and 7.3 (3.5 - 11.2)%, respectively. For both infections, reducing contacts between student grades within a school was found to be very effective and could be complementary to the main policies. Implementing strict quarantines on weekends were also found to be effective.

## Discussion

Outbreaks of acute respiratory infections such as influenza and the novel COVID-19 require an expansion of the available infection control policies. Here we report evidence in support of several such policies across outbreak scenarios and settings. For influenza, requiring isolation for fever is expected to reduce the typical attack rate by 29 (13 - 59)% and 70 (55 - 85)% with one and two days of post-fever isolation, respectively. This indicates that the CDC-recommended policy for schools, based on a single day of post-fever isolation, could be enhanced by requiring a second day of isolation. The result also holds in seasonal influenza in which vaccination is implemented. The isolation policy could be further strengthened by reducing contact between students during weekends. Using a shorter in-person school week (i.e., through longer weekends or remote learning) would also reduce the attack rate by 73 (64 - 88)% and 93 (91 - 97)% with four and three-day school weeks, respectively. The high percentage of reduction arises because these measures are expected to bring the epidemic under the outbreak threshold, and assumes no re-introduction of the infection from outside the school.

While we expected symptom-based isolation to be ineffective for COVID-19 since children are commonly asymptomatic^44^, it can still help in reducing outbreaks. Many authorities, including the CDC and WHO, recommend that those with symptoms isolate for a minimum of 10 days after symptom onset^11,12^. Indeed, we found that a one-day post-fever isolation policy would reduce the attack rate in schools by 7 (5 - 14)%, and with 14 days of fever isolation we estimated that the attack rate would change by 14 (5 - 26)%. The result shows that symptom-based isolation cannot be relied upon as a central policy in outbreak control for COVID-19, but it is not futile. Current CDC and WHO isolation policies for COVID-19 are expected to have effectiveness between the one-day and 14-day fever-based isolation policies above, but require only one day of isolation following any fever (CDC) and three days of isolation following cessation of symptoms (WHO). We found that shortening the school week does reduce the total number infected in an outbreak by 57 (52 - 64)% and 81 (79 - 83)%, with four and three days of in-person schooling, respectively.

Generally speaking, policies that isolate the infected, of which symptom-based isolation is a sub-type, are more preferable to closures. Unlike closures, symptom-based isolation allows healthy people in the community to continue living their lives uninhibited and reduces the considerable societal cost of school closures^45^. Consequently, a standing policy of isolation of infected individuals could potentially be sustained indefinitely. Shorter school weeks do affect all students, but they are less disruptive than full closures. They can also be maintained for extended times during the peak of the outbreak season, particularly if school days are replaced with remote learning (self study or e-learning). In severe outbreaks, a combination of policies would provide the best outcomes. High compliance by families would be critical to ensuring that the students are strongly isolated when the school is closed, and therefore families need to be supported.

While our model is driven by virological data and calibrated to several outbreaks, a few limitations are inherent in our approach. First, the effect of any policy depends on the context where it would be applied. The details of the school or institution would matter, and therefore, we provide an online version of the model, which can be calibrated for multiple situations. It may be possible to apply our findings to school-like contexts such as workplaces, prisons, or even the broader community, but such settings have significant features that may confound our findings. Lastly, symptom parameter information is based on average values for the population and it is expected that inter-individual and demographic variability might have some effect on outcomes. Future studies should attempt to evaluate isolation policies with agent-based models (e.g. ^15,46^) that can capture inter-individual variability in health trajectories and the network structure of the population^47,48^. Despite these limitations, our model captures essential aspects of acute respiratory outbreaks including progression through stages, the population structure and symptom trajectories.

In conclusion, in this study we have created a model of transmission of respiratory infection and considered the effects of two isolation policies. We confirmed that symptom-based policies would be effective in controlling influenza in a variety of scenarios. Furthermore for influenza outbreaks, we recommend that isolation is maintained for at least two days following the last day of fever. For both influenza and COVID-19 we found that using a shortened school-week of four days instead of five days could be effective in reducing the attack rate, and additional days would increase the effect. Policymakers tackling the influenza and COVID-19 outbreaks should consider implementing these policies for controlling outbreaks in schools and other settings.

## Data Availability

Complete source code and data are available

https://github.com/sashagutfraind/feverfighter/

## Supplementary Data

Supplemental Digital Content - are available online and include the model source code, an interactive online dashboard, and additional simulation data.

## Acknowledgements

Mark Dworkin, MD consulted on school absenteeism policies, and the following people provided comments: Alisa Ungar-Sargon, Michael Genkin, Michael Z. Levy, Edward A. Belongia, Sami Alhamdi, and Mitch Croal.

