## Supplementary material for "Effectiveness of Isolation Policies in Schools: Evidence from a Mathematical Model of Influenza and COVID-19": Methods Appendix

### Supplementary Article S1 - Expanded Methods

The text below describes the modeling methodology used in the study. In addition to this text, we provide the source code for the model, the associated data, and the expanded results (https://github.com/sashagutfraind/feverfighter). A live configurable dashboard for applying the model is also available (https://epi1.shinyapps.io/FeverFighter/).

**Modeling Methodology**
We use the methodology of a multi-stage SEIR model[1,2], that uses daily time steps combined with viral shedding and symptom data for influenza and COVID-19. In order to model the disease precisely, the model stratifies the infected population along three dimensions: the student cohort, the degree of isolation, and the stage of disease (Table S1 and Eqn A.1). The model divides the student population into cohorts representing the school grades (the school staff and the community could also be represented as their own cohort(s)). Each cohort is indexed by $i$ and includes its own variables: the susceptibles, $S_i$, infected (explained below), and recovered, $R_i$. Contact rates across cohorts and within them are described by a contact matrix, giving the model the ability to simulate a diverse set of institutions and communities. The model further divides the cohorts into isolated and unisolated ($H$ and $I$).

The model generalizes the classical SEIR model in which there is a sharp division between the exposed but not infectious population, and the infectious population (the E and the I in SEIR). In our model, each stage of the infection has a certain rate of infectiousness and symptoms (which can be zero, for the classical E compartment).

**Table S1. Variables of the models. Index $i$ indicates the cohort and $d$ the day of infection (up to 9 and 32 for influenza and COVID-19, respectively)**

| Variable | Interpretation |
|---|---|
| $S_i$ | Naïve persons in cohort $i$ (for immunity, $\tilde{S}_i(0) = S_i(0)(1 - vv_e)$, see below) |
| $I_{i,d}$ | Infected persons in cohort $i$ on day $d$ of infection who are not isolated |
| $H_{i,d}$ | Similarly, but who are *isolated* (e.g. at home) |
| $R_i$ | Recovered in cohort $i$ |

More precisely, let the force of infection $\lambda_i$ experienced by persons in cohort $i$ depend on the number of infected at each stage, the rate of viral shedding ($s_d$), and the contact matrix between all the cohorts $(c_{i,j})$. See Eqn A.1.

**Eqn A.1**. Our modeling framework - the multi-state discrete time SEIR infection model stratified by cohort and day of infection. Variables are dependent on time $t$. Return rates from isolation at day $r_d(p)$ depend on symptom isolation policy $p$, $\delta_{1,d}$ is the Kronecker delta function ($= 1$ if d is 1, and otherwise zero). $d_f$ is the final stage of infection. The parameters are listed in Table S2.

$$\Delta S_i = -bS_i\lambda_i$$
$$H_{i,d} = (1 - r_d(p))(H_{i,d-1} + I_{i,d-1})(1 - \delta_{1,d})$$
$$I_{i,d} = r_d(p)(H_{i,d-1} + I_{i,d-1})(1 - \delta_{1,d}) + \delta_{1,d}bS_i\lambda_i$$
$$\Delta R_i = H_{i,d_f} + I_{i,d_f}$$

$$\lambda_i = \sum_j \sum_d c_{i,j} s_d I_{j,d}$$

We model vaccination as a reduction in the initially susceptible population for each cohort, which depends on the vaccination rate and the vaccine efficacy: $\tilde{S}_i(0) = S_i(0)(1 - vv_e)$ . This transformation could be used to account for any resistance to infection, whether induced, or arising from genetics or prior exposure to similar pathogens.

Typical outbreak curves are shown in **Figure S1**.

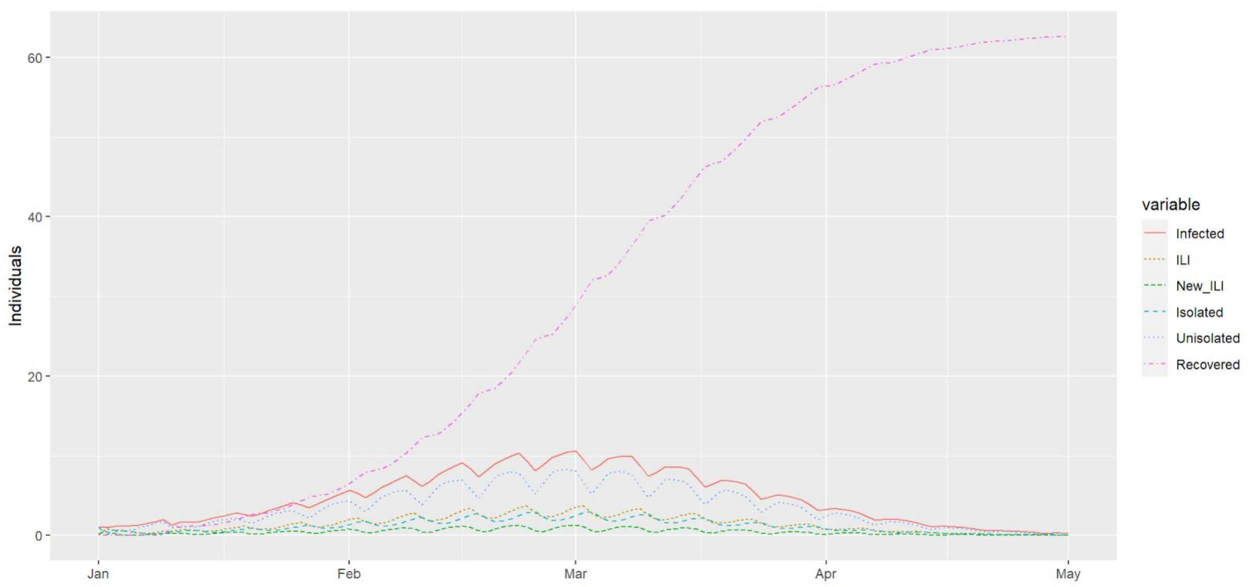

**Figure S1**. The forecasted epidemic curve of an influenza outbreak in a median school size based on US Department of Education data. Simulation is for the case with no formal isolation policy (where isolation occurs due to voluntary choice alone). Transmission is reduced during weekends, resulting in visible ripples in the outbreak curves every 7 days. ILI, influenza-like illness.

**Contact Rates**

We assumed that persons have the most physical contact with others in their cohort. The rate of contact with other cohorts is controlled by a parameter which was varied in our sensitivity analysis. To adjust for generally higher contact rates during the winter months, we use a seasonal term cf.[3,4] which multiplies the baseline contact rate, $b_0$ , by a factor that peaks on January 1st. :

$$b = b_0(1 + b_S cos(2\pi D/365)) \quad \text{Eqn A.2}$$

Here, D is the day of the year counting from January 1st. The value $b$ is multiplied by $b_h$ if the day falls on a weekend or by $b_c$ if the school is in closure or vacation (see Table S2).

**Sensitivity Analysis**

In sensitivity analysis, we assigned the parameters to truncated normal distributions with the mean, standard deviation (SD) and range obtained from previous research or calibration. For influenza, we then adjusted the transmission parameter of the model to give a 25% attack rate (thus matching the typical rate [5], after accounting for symptomatic rate[6]). For COVID-19, we adjusted the transmission parameter to give an 11.3% attack rate observed in a representative school outbreak (see Calibration section below). Experiments with alternative attack rates give qualitatively similar results (results not shown).

**Table S2.** Summary of parameters and their interpretation. All time rate constants are in units of day $^{-1}$. Parameters were obtained from previous studies or, when not available, by calibrating the model to an outbreak (see calibration section below). SD, standard deviation; CI, confidence interval.

| **Parameter** | **Interpretation** | **Range (SD)** | **Sources, if available** |
|---|---|---|---|
| $b_0$ | Core transmission rate | Influenza: 0.000402 (0) COVID-19: 0.000511 (0) | Calibrated. See section on model calibration |
| $b_h$ | Relative contact rate during weekends | 0.28014 (0.00875) | Calibrated. Comparable to school closure in [7] |
| $b_c$ | Relative contact rate during closures, holidays or vacations | 0.141479 (0.001822) | Calibrated. Comparable to school closure in [7] |
| $b_s$ | Relative seasonal amplitude of transmission | 0.478733 (0.001187) | Calibrated |
| $c_{i,j}$ | Relative contact rate for cohorts $i$ and $j$ where | 1 if $i = j$ else0.605565 (0.000672) | Calibrated. In some settings, this could be estimated from the degree of epidemiological separations of the cohorts |
| $l$ | Symptomatic rate | Influenza: 84% (CI: 81-87%) COVID-19: 18.1% (CI:13.9-22.9) | For influenza, based on meta-analysis of asymptomatic rate[6]; for COVID-19[8] |
| $p$ | Fraction of persons complying with isolation | 0.158385 (0.199545) | Calibrated. Quantifies compliance with the policy |
| $r_d$ | Fraction of persons returning to cohort on day $d$ after infection | Based on symptom data | See Table S3 and Table S4 below |
| $s_d$ | Shedding at day d | Table S3 and Table S4 | |
| $t_{start}$ | Day of first infected case | Input data or calibration | Often reported approximately in post-outbreak investigations |
| $v$ | Vaccination rate (if vaccine available) | 0.8 (0.1) | Seasonal flu: 60% [0.5, 0.7][9] Novel flu strains: 0% |
| $v_e$ | Vaccine efficacy (if vaccine available) | Influenza 0.5 (0.1) Covid-19: 0.7 (0.2) | For influenza[10,11]. Higher mean for COVID-19 based on novel mRNA vaccines. For novel infections with no immune response, 0%. |
| $x_i$ | Persons in cohort $i$ | Normal: 70 Large: 140 | For a typical K-5 for the US[12]. Six cohorts per school[13] |
| $y$ | Fever attention | 0.698836 (0.001567) | Calibrated. Fraction of persons who would return from home, of those having symptoms on the previous day |

To estimate the rate of symptomatic COVID-19 cases, our priority was to use only studies that had unbiased sampling of all at-risk persons, since surveillance-based methods tend to capture individuals with higher expression of symptoms[6]. Therefore, we adopted the estimate of Poletti et al.[8] which tested all household members of known COVID-19 cases and is unique in reporting the symptom rate for ages 0 to 19, instead of all age ranges. It used a symptom definition of upper or lower respiratory tract symptoms, or fever ≥37.5 °C and a sample size of N=692.

Their estimate is consistent with a large random survey from Spain[14] that reported a rate of 34.53% but that included adults that have a higher rate of symptomatic infections[8,15].

**Modeling of isolation behaviors and the effect of isolation policy**
We model the complex interaction between the factors that affect the population. As quantified below, the rate of returning to school is decreased at the more severe stages of the disease, when the disease has a higher symptomatic rate, or when the patients are more aware of their symptoms. As described below, the symptom-based isolation policy is modeled as a decrease in the rate at which students return to school.

To be exact, let the values $r_d$ express the fraction of persons returning from home on day $d$ after they become infected when no policy is in place. It is modeled as a function of four factors: the symptoms in the previous days $(f_s)$, the symptom propensity (i.e. rate of symptomatic infections) $l \in [0, 1]$, the attention to symptoms $y \in [0, 1]$, and the compliance to policy (if any) $p \in [0, 1]$. To be precise, in the simplest case (no isolation policy), increasing $f_{d-1}$, $l$ or $y$ would decrease the return rate: $r_d = 1 - lyf_{d-1}$. The introduction of the isolation policy decreases the return to school rate at day $d$ by making the persons more attentive to the recent days of symptoms. Namely, under a one-day isolation policy, the rate is modified to:

$$r_d = 1 - [l(y + (1 - y)p)] f_{d-1} \text{ on day } d; \text{ Eqn A.3}$$

This model ensures that the rate of symptoms $lf_{d-1}$ sets an upper bound on the effectiveness of the symptom-isolation policy. In the case of 100% compliance and 100% symptom attention, the return rate is $1 - lf_{d-1}$ and not lower. Under a two-day isolation policy, the rate on day $d$ is given by replacing $f_{d-1}$ in Eqn A.3 with $max(f_{d-1}, f_{d-2})$, and in general, $max_{i=1..d-1}(f_i)$ for longer isolation policies (see Table S3 and Table S4).

**Viral Shedding and Symptom Burden**
Influenza viral shedding appears to vary by subtype. However, the patterns of shedding were similar in both children and adults[16,17], and between the seasonal and p(H1N1) outbreaks [17–19]. We allowed for a proportion $l$ of the infections to be asymptomatic[16,20], which in our model increases the rates of return to the community and transmission, while also reducing the effectiveness of control policies. Meta-analysis of influenza studies[21] was used to determine shedding and symptom rates by disease state (see Table S3, Table S4).

**Table S3.** Daily symptom and shedding rates for influenza from [21]. Viral shedding rate is based on log10 of titers, and is multiplied by the transmissibility parameter in the model. Symptom scores are relative to their peak. New influenza-like illness (ILI) rates is the probability of newly reporting ILI on a given day. Predicted return rates when symptom propensity is $l = 0.84$ and symptom attention is $y = 0.5$. Return rates: A = no policy, B = policy of one day of isolation with 0.5 compliance, C = policy of one day of isolation with 100% compliance. As the rate of compliance rises, the return rate generally decreases since students self-isolate at home. Rows are days from the time of infection. Total Score is an overall indicator of illness reported in [21].

| | | Symptoms | | | | | Return rate by scenario | | |
|---|---|---|---|---|---|---|---|---|---|
| Day after infection | Viral Shedding | Total Score | Fever (Systemic) | Respiratory | Nasal | New ILI Rates | A | B | C |
| 1 | 1.89 | 0.25 | 0.12 | 0.15 | 0.18 | 0.25 | 1.00 | 1.00 | 1.00 |

| | | | | | | | | | |
|---|---|---|---|---|---|---|---|---|---|
| 2 | 3.00 | 0.67 | 0.93 | 0.66 | 0.79 | 0.50 | 0.919 | 0.911 | 0.903 |
| 3 | 2.63 | 0.85 | 0.75 | 0.95 | 0.94 | 0.25 | 0.347 | 0.281 | 0.215 |
| 4 | 2.16 | 0.67 | 0.60 | 0.91 | 0.91 | 0.00 | 0.478 | 0.425 | 0.372 |
| 5 | 1.54 | 0.47 | 0.30 | 0.56 | 0.70 | 0.00 | 0.58 | 0.538 | 0.496 |
| 6 | 1.07 | 0.18 | 0.17 | 0.56 | 0.47 | 0.00 | 0.789 | 0.767 | 0.746 |
| 7 | 0.74 | 0.06 | 0.08 | 0.49 | 0.17 | 0.00 | 0.881 | 0.869 | 0.857 |
| 8 | 0.30 | 0.06 | 0.07 | 0.36 | 0.02 | 0.00 | 0.941 | 0.935 | 0.929 |
| 9 | 0.35 | 0.00 | 0.08 | 0.14 | 0.00 | 0.00 | 0.951 | 0.946 | 0.941 |

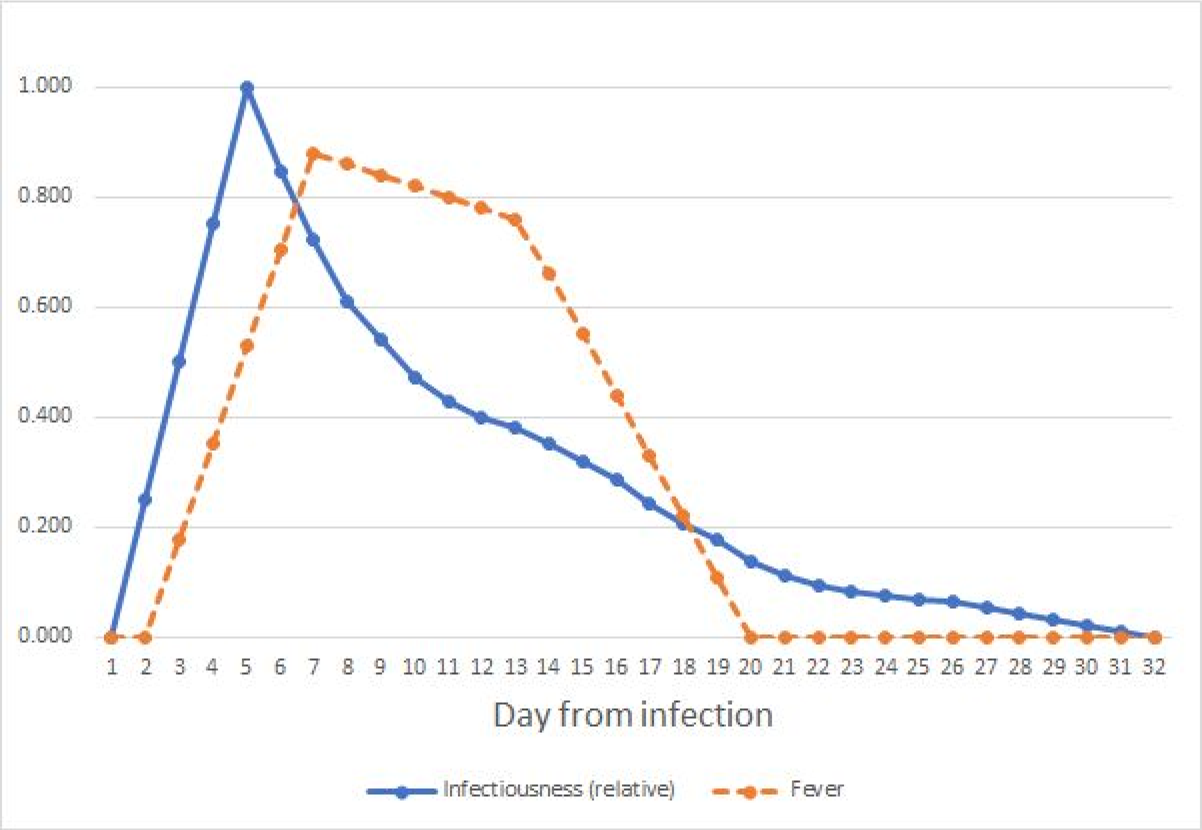

**Figure S2**. Estimated SARS-CoV-2 infectiousness and symptom propensity in symptomatic cases. Infectiousness is based on [22] with linear interpolation added before day 5 and after day 28[23–27]. Fever scores for patients with fever are based on collections of case reports[28].

**Table S4.** Estimated symptoms, shedding rates, and return rates for symptomatic persons infected with SARS-CoV-2. Asymptomatic infections are accounted for through a separate parameter. Return rates calculated for when attention is focused on fever, symptom propensity rate is $l = 0.1809$, and symptom attention is $y = 0.5$. Return rates: A = no policy, B = policy of one day of isolation with 0.5 compliance, C = policy of one day of isolation with 100% compliance. As the rate of compliance rises, the return rate generally decreases since students self-isolate at home. Rows are days from the time of infection.

| | | **Symptoms** | **Return rate by scenario** | | | | | | **Symp toms** | **Return rate by scenario** | | |
|---|---|---|---|---|---|---|---|---|---|---|---|---|
| **Day after infection** | **Viral Shedding** | **Fever (Systemic)** | **A** | **B** | **C** | | **Day after infection** | **Viral Shed ding** | **Fever (Syste mic)** | **A** | **B** | **C** |
| **1** | 0.000 | 0 | 1 | 1 | 1 | | **17** | 0.244 | 0.330 | 0.934 | 0.927 | 0.92 |
| **2** | 0.250 | 0 | 1 | 1 | 1 | | **18** | 0.207 | 0.220 | 0.95 | 0.945 | 0.94 |
| **3** | 0.500 | 0.176 | 1 | 1 | 1 | | **19** | 0.177 | 0.110 | 0.967 | 0.964 | 0.96 |
| **4** | 0.750 | 0.352 | 0.973 | 0.971 | 0.968 | | **20** | 0.139 | 0 | 0.983 | 0.982 | 0.98 |
| **5** | 1.000 | 0.528 | 0.947 | 0.942 | 0.936 | | **21** | 0.112 | 0 | 1 | 1 | 1 |
| **6** | 0.845 | 0.704 | 0.92 | 0.912 | 0.904 | | **22** | 0.095 | 0 | 1 | 1 | 1 |
| **7** | 0.721 | 0.880 | 0.894 | 0.883 | 0.873 | | **23** | 0.082 | 0 | 1 | 1 | 1 |
| **8** | 0.611 | 0.860 | 0.867 | 0.854 | 0.841 | | **24** | 0.077 | 0 | 1 | 1 | 1 |
| **9** | 0.540 | 0.840 | 0.871 | 0.857 | 0.844 | | **25** | 0.070 | 0 | 1 | 1 | 1 |
| **10** | 0.473 | 0.820 | 0.874 | 0.861 | 0.848 | | **26** | 0.067 | 0 | 1 | 1 | 1 |
| **11** | 0.427 | 0.800 | 0.877 | 0.864 | 0.852 | | **27** | 0.056 | 0 | 1 | 1 | 1 |
| **12** | 0.400 | 0.780 | 0.88 | 0.867 | 0.855 | | **28** | 0.044 | 0 | 1 | 1 | 1 |

| | | | | | | | | | | | | |
|---|---|---|---|---|---|---|---|---|---|---|---|---|
| **13** | 0.379 | 0.760 | 0.883 | 0.871 | 0.859 | | **29** | 0.033 | 0 | 1 | 1 | 1 |
| **14** | 0.351 | 0.660 | 0.886 | 0.874 | 0.863 | | **30** | 0.022 | 0 | 1 | 1 | 1 |
| **15** | 0.318 | 0.550 | 0.901 | 0.891 | 0.881 | | **31** | 0.011 | 0 | 1 | 1 | 1 |
| **16** | 0.287 | 0.440 | 0.917 | 0.909 | 0.901 | | **32** | 0.000 | 0 | 1 | 1 | 1 |

**Model Validation and Calibration**

The model has been validated by comparing it to empirical data on three influenza outbreaks and one outbreak of COVID-19. The available data generally indicates the daily number of new ILI cases or the daily number of new absences, and both were also calculated from the model. In all calibrations, we found the model to give a tight fit to the data matching the attack rate, the peak date, and the overall shape, as illustrated in **Figure S3, Figure S4, and Figure S5**. For COVID-19, the fit is reported in Table S5.

For validation, we needed to estimate from the model the number of individuals with new influenza-like illness (ILI) and the number of newly absent. For ILI, the number of individuals with ILI at a given time $t$ is computed by considering $J_s(t)$ - the number of infected individuals on date $t$ (including isolated and unisolated) who are in stage $s$ and the rate of new ILI cases at stage $s$, $l_s$. The quantity is multiplied by $l_s$ - the fraction of individuals that develop ILI (set at 84%[20]): $ILI(t) = l \sum_s l_s J_s(t)$ .

The symptom data indicates that ILI symptoms first arise 24 hours after influenza infection and peak around 2.5 days after infection. Accordingly, we assume that, of the population reporting ILI symptoms, ¼, ½, and ¼ first reports on day 1, 2, and 3, respectively.

We calculated the new absentees per day by considering the number of infected and the return rate. The number of isolated persons is given by: $\sum_s (1 - r_s) I_s(t)$. Furthermore, the number of newly-absent (i.e. isolated) students is given by $\sum_s \prod_{q=1}^{s-1} r_q (1 - r_s) I_s(t)$. We will use this formula in calibrating the model to absenteeism data below.

Unknown parameter values and ranges were estimated using a genetic algorithm[29]. The algorithm adjusted the values of the unknown parameters to match the well-characterized A(H1N1)v influenza outbreak in a boarding school[30]. Parameter ranges were estimated from the distribution of parameters in the top 10% of the solutions. A separate set of simulations with seasonal influenza calibrated to an outbreak of seasonal influenza [31]..

**(A)**

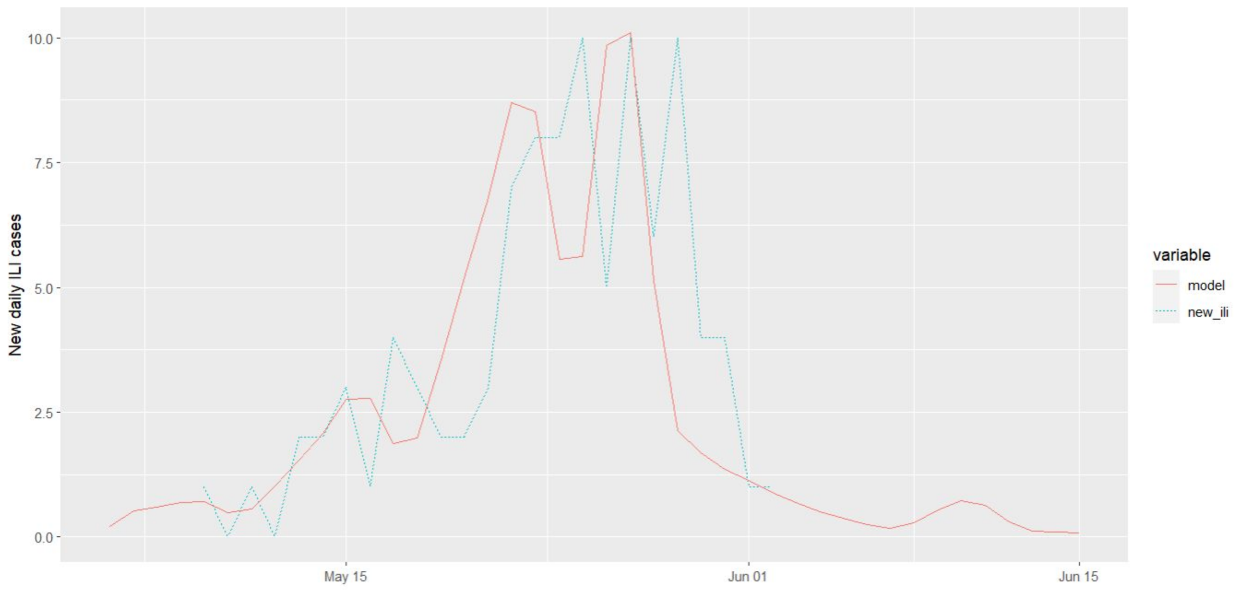

**(B)**

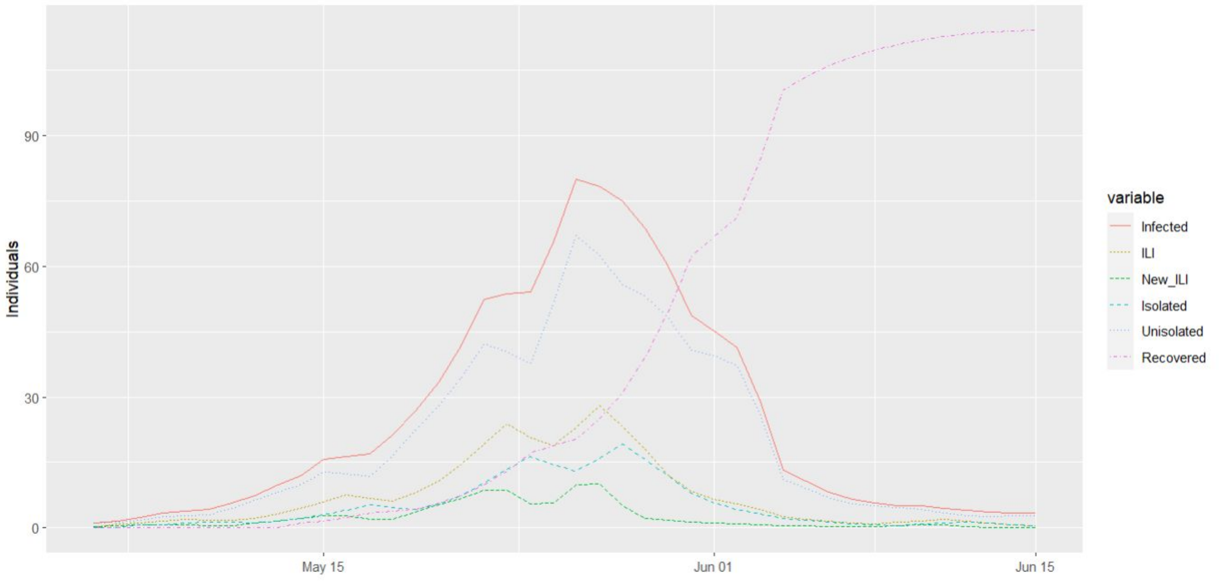

**Figure S3**. Calibration to data from a boarding school in South East England during the A(H1N1)v outbreak from May 9, 2009 through June 2, 2009, infecting 101 of the 1307 students[30]. (A) model predictions and actual new cases of influenza-like illness (ILI) and (B) estimated course of the epidemic from the model.

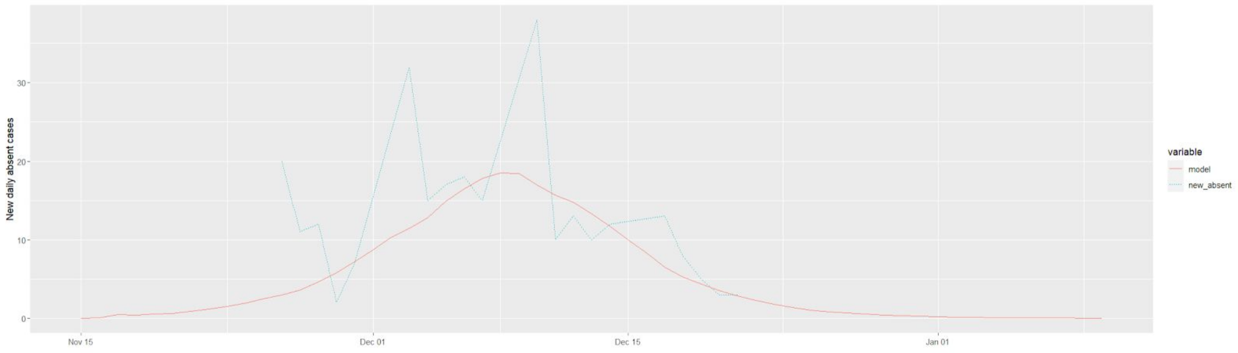

**(A)**

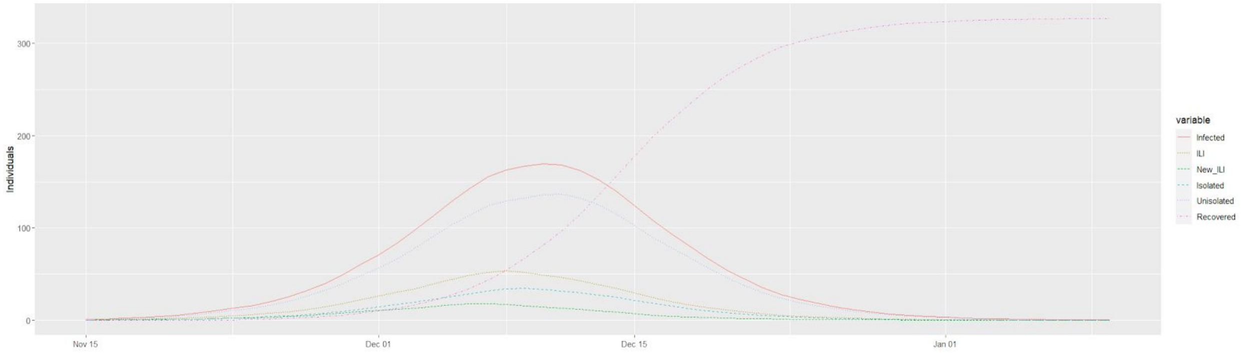

**(B)**

**Figure S4**. Calibration to absenteeism for a primary school in Thames Valley, UK during the 2012/2013 influenza season abstracted from [31]. The data shows spikes of new absences every Monday due to lack of data collection on Saturdays and Sundays. (A) model predictions and match to actual new cases of absence from school and (B) estimated course of the epidemic from the model.

**(A)**

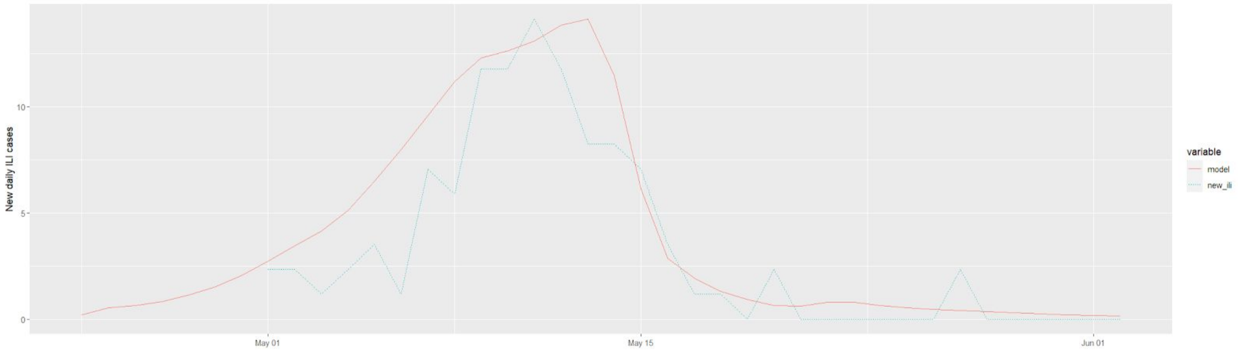

**(B)**

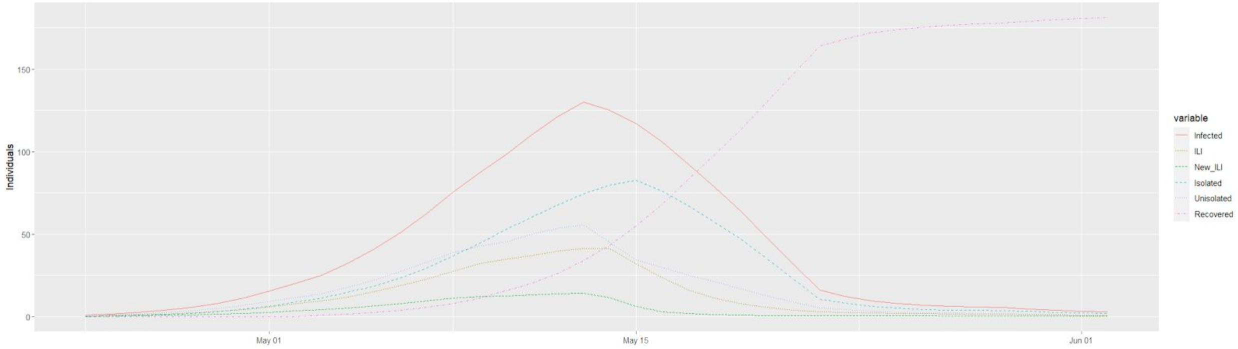

**Figure S5**. Calibration to data from an elementary school in Pennsylvania affected by the 2009 Pandemic Influenza A(H1N1)[32]. (A) model predictions and actual new cases of influenza-like illness (ILI) and (B) estimated course of the epidemic from the model.

**Table S5**: Outbreak of COVID-19 in the Rehavia Gymnasia (Jerusalem, Israel) - high school with grades 7-12 containing 1200 students and 180 staff. The school was closed starting 5/28 and complete serosurvey found 159 infections[33].

| Date | Reported new cases | Predicted Infected | Predicted daily new symptomatic | | Date | Reported new cases | Predicted Infected | Predicted daily new symptomatic |
|---|---|---|---|---|---|---|---|---|
| 5/11/2020 | | 1 | 0 | | 5/21/2020 | | 18 | 0 |
| 5/12/2020 | | 1 | 0 | | 5/22/2020 | | 19 | 0 |
| 5/13/2020 | | 1 | 0 | | 5/23/2020 | | 20 | 0 |
| 5/14/2020 | | 2 | 0 | | 5/24/2020 | | 34 | 0 |
| 5/15/2020 | | 2 | 0 | | 5/25/2020 | | 50 | 0 |
| 5/16/2020 | | 2 | 0 | | 5/26/2020 | 1st known case | 71 | 1 |
| 5/17/2020 | | 4 | 0 | | 5/27/2020 | 2 new cases | 98 | 1 |
| 5/18/2020 | | 6 | 0 | | 5/28/2020 | Last day of classes | 136 | 1 |
| 5/19/2020 | | 9 | 0 | | 5/29/2020 | School shut down | 142 | 2 |
| 5/20/2020 | | 12 | 0 | | Final serological survey of the school | Actual infected 159 | Predicted infected 159 | - |
